# Prognostic Value of Left Ventricular ^18^F-Florbetapir Uptake in Systemic Light-Chain Amyloidosis

**DOI:** 10.1101/2023.09.13.23295520

**Authors:** Olivier F. Clerc, Yesh Datar, Sarah AM Cuddy, Giada Bianchi, Alexandra Taylor, Dominik C Benz, Matthew Robertson, Marie Foley Kijewski, Michael Jerosch-Herold, Raymond Y Kwong, Frederick L Ruberg, Ronglih Liao, Marcelo F Di Carli, Rodney H Falk, Sharmila Dorbala

**Author notes:** Address for Correspondence Sharmila Dorbala, MD, MPH Cardiac Amyloidosis Program, Division of Cardiology, Department of Medicine Cardiovascular Imaging Program, Division of Cardiology and Department of Radiology Division of Nuclear Medicine and Molecular Imaging, Department of Radiology Brigham and Women’s Hospital and Harvard Medical School 75 Francis St, Boston, MA 02115, USA; Twitter Handle: @DorbalaSharmila.

## Abstract

**Background:** Myocardial immunoglobulin light-chain (AL) amyloid deposits trigger heart failure, cardiomyocyte stretch and myocardial injury, leading to adverse cardiac outcomes. Positron emission tomography/computed tomography (PET/CT) with ^18^F-florbetapir, a novel amyloid-targeting radiotracer, can quantify left ventricular (LV) amyloid burden, but its prognostic value is not known. Therefore, we aimed to evaluate the prognostic value of LV amyloid burden quantified by ^18^F-florbetapir PET/CT and to identify mechanistic pathways mediating its association with outcomes.

**Methods:** Eighty-one participants with newly-diagnosed systemic AL amyloidosis were prospectively enrolled and underwent ^18^F-florbetapir PET/CT. LV amyloid burden was quantified using ^18^F-florbetapir LV percent injected dose (%ID). Mayo AL stage was determined using troponin T, N-terminal pro-B-type natriuretic peptide (NT-proBNP), and difference between involved and uninvolved free light chain levels. Major adverse cardiac events (MACE) were defined as all-cause death, heart failure hospitalization, or cardiac transplantation within 12 months.

**Results:** Among participants (median age 61 years, 57% males), 36% experienced MACE. Incidence of MACE increased across tertiles of LV amyloid burden from 7% to 63% (p<0.001). LV amyloid burden was significantly associated with MACE in univariable analysis (hazard ratio 1.45, 95% confidence interval 1.15-1.82, p=0.002). However, this association became non-significant in multivariable analyses adjusted for Mayo AL stage. Mediation analysis showed that the association between ^18^F-florbetapir LV %ID and MACE was primarily mediated by NT-proBNP (p<0.001), a marker of cardiomyocyte stretch and component of Mayo AL stage.

**Conclusion:** In this first study to link cardiac ^18^F-florbetapir uptake to subsequent outcomes, LV amyloid burden estimated by LV %ID predicted MACE in AL amyloidosis. But this effect was not independent of Mayo AL stage. LV amyloid burden was associated with MACE primarily via NT-pro-BNP, a marker of cardiomyocyte stretch and component of Mayo AL stage. These findings provide novel insights into the mechanism through which myocardial AL amyloid leads to MACE.

**Clinical Perspective:** In systemic light-chain (AL) amyloidosis, cardiac involvement is the key determinant of adverse outcomes. Usually, prognosis is based on the Mayo AL stage, determined by troponin T, N-terminal pro-B-type natriuretic peptide (NT-proBNP), and the difference between involved and uninvolved immunoglobulin free light chain levels (dFLC). Cardiac amyloid burden is not considered in this staging. In the present study, we used the amyloid-specific radiotracer ^18^F-florbetapir to quantify left ventricular (LV) amyloid burden in 81 participants with newly-diagnosed AL amyloidosis and evaluated its prognostic value on major adverse outcomes (MACE: all-cause death, heart failure hospitalization, or cardiac transplantation within 12 months). We found that higher LV amyloid burden by ^18^F-florbetapir positron emission tomography/computed tomography (PET/CT) was strongly associated with MACE. However, this association became non-significant after adjustment for the Mayo AL stage. Mediation analysis offered novel pathophysiological insights, implying that LV amyloid burden leads to MACE predominantly through cardiomyocyte stretch and light chain toxicity (by NT-proBNP), rather than through myocardial injury (by troponin T), also considering the severity of plasma cell dyscrasia (by dFLC). This mediation by NT-proBNP may explain why the association with outcomes was non-significant with adjustment for Mayo AL stage. Together, these results establish quantitative ^18^F-florbetapir PET/CT as a valid method to predict adverse outcomes in AL amyloidosis. These results support the use of ^18^F-florbetapir PET/CT to measure the effects of novel fibril-depleting therapies, in addition to plasma cell therapy, to improve outcomes in systemic AL amyloidosis.

## Introduction

In systemic light-chain (AL) amyloidosis, clonal plasma cells produce a large quantity of circulating abnormal immunoglobulin light chains. These light chains are prone to misfolding, which leads to aggregation as amyloid fibrils in multiple organs, extracellular space expansion, organ dysfunction and death.^1,2^ Among affected organs, cardiac involvement is the most important determinant of adverse outcomes in AL amyloidosis.^1,2^ If untreated, AL amyloid cardiomyopathy is highly fatal, with a median survival of 6 months.^1^ Even with state-of-the-art therapies, including daratumumab, the 12-month mortality remains substantial, about 20%.^3,4^ The importance of cardiac involvement is reflected in the Mayo prognostic staging, which is mostly based on cardiac serum biomarkers: troponin T, N-terminal pro-B-type natriuretic peptide (NT-proBNP) and the difference between involved and uninvolved immunoglobulin free light chains (dFLC).^5^

Imaging metrics of cardiac AL amyloid burden are not included in the current prognostic staging.^6,7^ However, a strong prognostic value was demonstrated for cardiac imaging in AL amyloidosis, particularly for echocardiography with structural, diastolic and strain metrics,^8,9^ as well as for magnetic resonance imaging (MRI) with metrics of cardiac structure and function, late gadolinium enhancement, T1 mapping and extracellular volume.^8,10,11^ Furthermore, the novel, specific β-amyloid-binding radiotracers for positron emission tomography/computed tomography (PET/CT) have shown promise to diagnose and quantify cardiac AL amyloidosis.^12–18^ Moreover, cardiac uptake of the radiotracer ^11^C-Pittsburgh compound B has been associated with all-cause death in AL amyloidosis.^19,20^ However, whether metrics of amyloid burden derived from β-amyloid-binding radiotracer PET/CT add prognostic value over the Mayo stage, and whether their associations with outcomes are direct or indirect, i.e., through specific pathophysiological pathways, are unknown.

Among amyloid-specific radiotracers for PET/CT, ^18^F-florbetapir exhibits a high cardiac uptake in AL amyloidosis,^13^ and can detect cardiac AL amyloid deposits even in participants with normal cardiac serum biomarkers (troponin T, NT-proBNP) and left ventricular (LV) thickness.^16^ This suggests the potential to quantify cardiac amyloid burden and thereby predict outcomes in AL amyloidosis. Therefore, the aims of this study of participants with new diagnosis of AL amyloidosis with or without known cardiac involvement were: 1) to assess the prognostic value of cardiac amyloid burden metrics derived from ^18^F-florbetapir PET/CT images for major adverse cardiac events (MACE), 2) to evaluate the independent prognostic value of cardiac amyloid burden metrics compared with the Mayo stage, and 3) to investigate mechanistic pathways of the effects of cardiac amyloid burden on MACE using mediation analysis. We also explored all-cause death as a secondary endpoint.

## Methods

### Participant Selection Criteria

This prospective study was approved by the Mass General Brigham Human Research Committee, and each participant provided written informed consent. From 2016 to 2022, participants with systemic AL amyloidosis were recruited primarily from Brigham and Women’s Hospital (Boston), as well as form Dana-Farber Cancer Institute (Boston), Massachusetts General Hospital (Boston), Boston Medical Center, Boston University School of Medicine, and Memorial Sloan Kettering Cancer Center (New York) into the Molecular Imaging of Primary Amyloid Cardiomyopathy cohort study (MICA; NCT02641145).^16,21–24^ Systemic AL amyloidosis was diagnosed by standard criteria, with confirmation of the plasma cell dyscrasia in the bone marrow, as well as endomyocardial or systemic biopsy with determination of amyloid type by immunohistochemistry or mass spectrometry.^25^ Key exclusion criteria were significant non-amyloid cardiac disease (e.g., coronary artery disease, severe valvular disease), cardiac pacemaker, implanted cardioverter-defibrillator, estimated glomerular filtration rate (eGFR) <30 mL/min/1.73m^2^ and severe claustrophobia. All participants from the MICA study at therapy initiation for newly-diagnosed systemic AL amyloidosis were included (N=81), while participants in remission after therapy were excluded (N=25). Treatment decisions for AL amyloidosis were made by the patients’ primary hematologist without knowledge of the results of the PET/CT study.

### Laboratory Evaluation and Mayo AL Prognostic Staging

Serum troponin T, NT-proBNP, free light chain levels, creatinine and hemoglobin were measured at Brigham and Women’s Hospital central laboratory. The eGFR was calculated using the Chronic Kidney Disease Epidemiology Collaboration (CKD-EPI) formula. The Mayo AL stage was calculated starting at one point, with one more point added for each positive criterion: troponin T ≥0.025 ng/mL, NT-proBNP ≥1800 pg/mL and dFLC ≥180 mg/L.^5^

### 18F-Florbetapir PET/CT Acquisition

All study participants underwent a cardiac PET/CT (Discovery STE, Discovery RX, or Discovery MI, GE Healthcare, Chicago, IL, USA) with 30-min list-mode acquisition at the Brigham and Women’s Hospital. A low-dose chest CT scan was acquired for patient positioning and attenuation correction of the PET emission data. ^18^F-florbetapir was injected intravenously 1 minute after the beginning of PET acquisition. The median injected ^18^F-florbetapir activity was 9.0 mCi (interquartile range [IQR] 8.5‒9.9), with a median effective dose of 6.9 mSv (IQR 6.5‒7.5), including 0.5 mSv for the low-dose CT scan. Static images of the heart were reconstructed using data from 4 minutes to 30 minutes after radiotracer injection, based on previously validated data on ^18^F-florbetapir washout from the blood pool.^13^

### 18F-Florbetapir PET/CT Quantitation of Amyloid Burden

LV ^18^F-florbetapir activity concentration was measured volumetrically on deidentified static images using PMOD software (PMOD Technologies LLC, Zurich, Switzerland). A volume of interest (VOI) was placed on PET images using fused CT images to define the LV contours, including the blood pool and the interventricular septum. To minimize observer bias in myocardial tracings, we used the iso-contouring function from PMOD on the LV tracings to delineate the myocardial wall uptake without blood pool as LV wall VOI. This was performed by automatic thresholding of VOI activity concentration above two times the mean blood pool activity concentration in a 10-mm spherical left atrial VOI, as previously done.^13,16^ To quantify cardiac amyloid burden, we used the LV percent injected dose (%ID), calculated as the LV wall VOI mean activity concentration multiplied by the LV wall VOI volume and divided by the injected activity. All activity values were decay-corrected to scan start. LV %ID quantifies the total amount radiotracer bound to the LV myocardium, and therefore, the LV amyloid burden. Other uptake metrics were considered, but LV %ID was chosen for its ability to measure the total amyloid burden and its superior prognostic ability (<u>Supplemental Methods</u>, Supplemental Table 1).

### Echocardiography

All participants underwent echocardiography, but only exams performed within 6 months of PET/CT were included (median absolute difference 5 days, IQR 1–27, 6 exams excluded). Measurements were performed using dedicated software (TOMTEC Image Arena version 4.6, TOMTEC Imaging Systems GmbH, Unterschleissheim, Germany).

### Study Endpoints and Event Ascertainment

MACE was defined as a composite of all-cause death, heart failure hospitalization, or cardiac transplantation. Event ascertainment was performed by phone calls to participants and review of electronic medical records, with 99% successful follow-up. Heart failure hospitalization was adjudicated blinded to imaging results. The primary endpoint for this study was MACE during the first 12 months to maintain a close association with the diagnosis of AL amyloidosis. The secondary endpoint was all-cause death within 12 months. Sensitivity analyses using adverse events during the complete follow-up were performed (<u>Supplemental Materials</u>).

### Statistical Analysis

Continuous variables were summarized as median with IQR, and categorical variables were as frequency with percentage. They were analyzed by tertiles of LV amyloid burden based on ^18^F-florbetapir PET/CT LV %ID. The Jonckheere-Terpstra test or the Cochran-Armitage test were used to calculate p-values for trend for continuous or categorical variables, respectively. Kaplan-Meier analysis was conducted by tertiles of LV amyloid burden with the log-rank test. Cox regression was performed to evaluate associations with adverse outcomes. Proportional hazards assumptions were checked. Finally, we performed a causal mediation analysis of the association between LV amyloid burden and MACE,^26,27^ using the biomarkers composing the Mayo stage, which have been associated with adverse outcomes.^5^ We assumed that NT-proBNP (reflecting cardiomyocyte stretch and light chain toxicity) and troponin T (reflecting myocardial injury) were potential mediators, while dFLC (reflecting circulating light chains) was a potential confounder of the association between LV amyloid burden and MACE.^28^ The choice of mediator or confounder was based on the direction of the assumed causality: LV amyloid burden (%ID) causes cardiomyocyte stretch (NT-proBNP) and myocardial damage (troponin T), but circulating light chains (dFLC) cause LV amyloid burden, while cardiomyocyte stretch, myocardial damage and circulating light chains cause MACE. Thus, LV amyloid burden can determine MACE directly, or indirectly through a mediation by cardiomyocyte stretch and myocardial damage, and the association between LV amyloid burden and MACE may be affected by confounding from circulating light chains. Linear regression was used to evaluate associations between ^18^F-florbetapir uptake and biomarkers, log-transformed to comply with assumptions for linear regression. An accelerated failure time model (parametric survival regression) with log-normal distribution was used to study the association between ^18^F-florbetapir uptake and MACE with adjustment for biomarkers in the mediation analysis, because the rare outcome assumption allowing the use of Cox regression was not met.^26,27^ The log-normal distribution exhibited a better fit than other potential distributions and was therefore chosen. Given the continuous nature of LV amyloid burden, we specified the treatment value as the median of the highest tertile of LV %ID, and the control value as the median of the lowest tertile of LV %ID. For the mediation analysis, confidence intervals and p-values were computed using bootstrapping with 1000 draws and percentile confidence intervals.^27^ Two-sided p-values were reported and considered statistically significant if <0.05. Data were analyzed using R version 4.3.1 (R Core Team. R: A language and environment for statistical computing. R Foundation for Statistical Computing, Vienna, Austria) and the packages *tidyverse*, *DescTools*, *gtsummary*, *rstatix*, *survival*, *survminer, ggsurvfit* and *mediation*. The manuscript was written according to the STrengthening the Reporting of OBservational studies in Epidemiology (STROBE) and A Guideline for Reporting Mediation Analyses (AGReMA) statements.^29,30^

## Results

### Participant Characteristics at Baseline

In our 81 participants with newly-diagnosed AL amyloidosis, the median age was 61 years (IQR 57–67), 46 were males (57%), and 61 had cardiac involvement by elevated serum cardiac biomarkers (75%). Increasing tertiles of LV amyloid burden by ^18^F-florbetapir %ID were associated with lower systolic blood pressure, higher serum cardiac biomarkers, higher Mayo AL stage and worse LV global longitudinal strain (GLS) on echocardiography (all p<0.001; Table 1). This suggests that LV amyloid burden assessed by ^18^F-florbetapir %ID quantifies the functional severity of AL cardiomyopathy (Figure 1). Among participants, 73% were treated with a cyclophosphamide-bortezomib-dexamethasone (CyBorD) regimen, 52% with daratumumab, 43% received other chemotherapeutic agents, and 21% underwent autologous stem cell transplantation (ASCT; Table 1).

**Figure 1.**
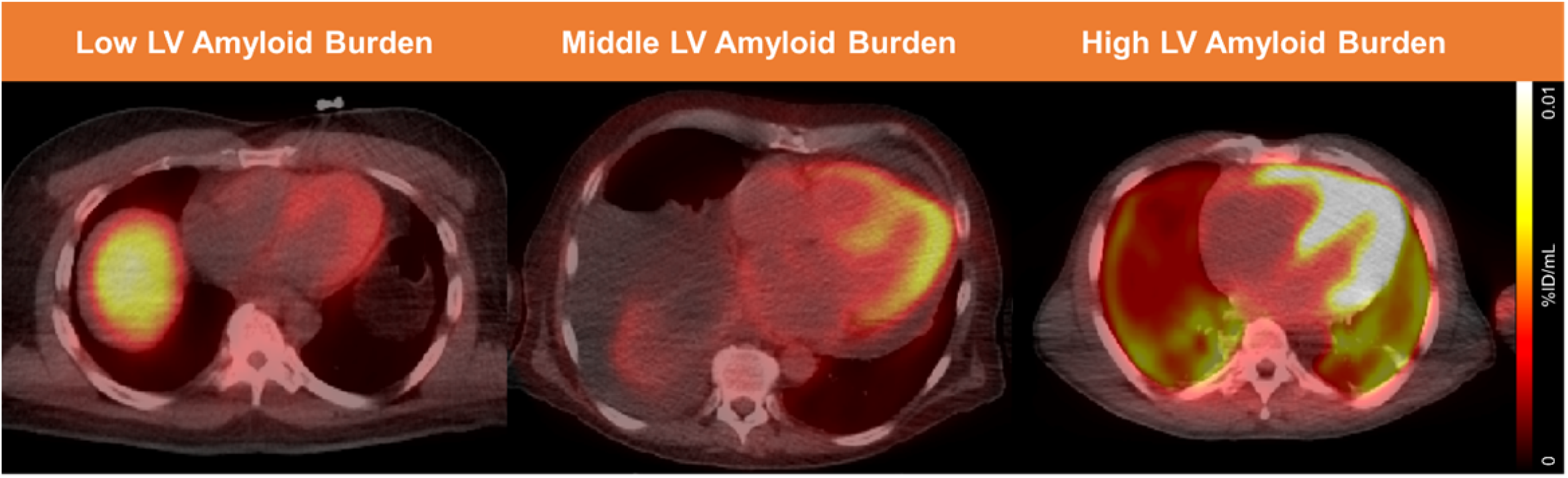
Cardiac Amyloid Quantification with ^18^F-Florbetapir PET/CT. Images from ^18^F-florbetapir PET/CT with low, middle and high LV amyloid burden are presented, showing clear differences in left ventricular uptake. %ID/mL: Percent injected dose by milliliter. LV: Left ventricular. PET/CT: Positron emission tomography/computed tomography.

**Table 1.**
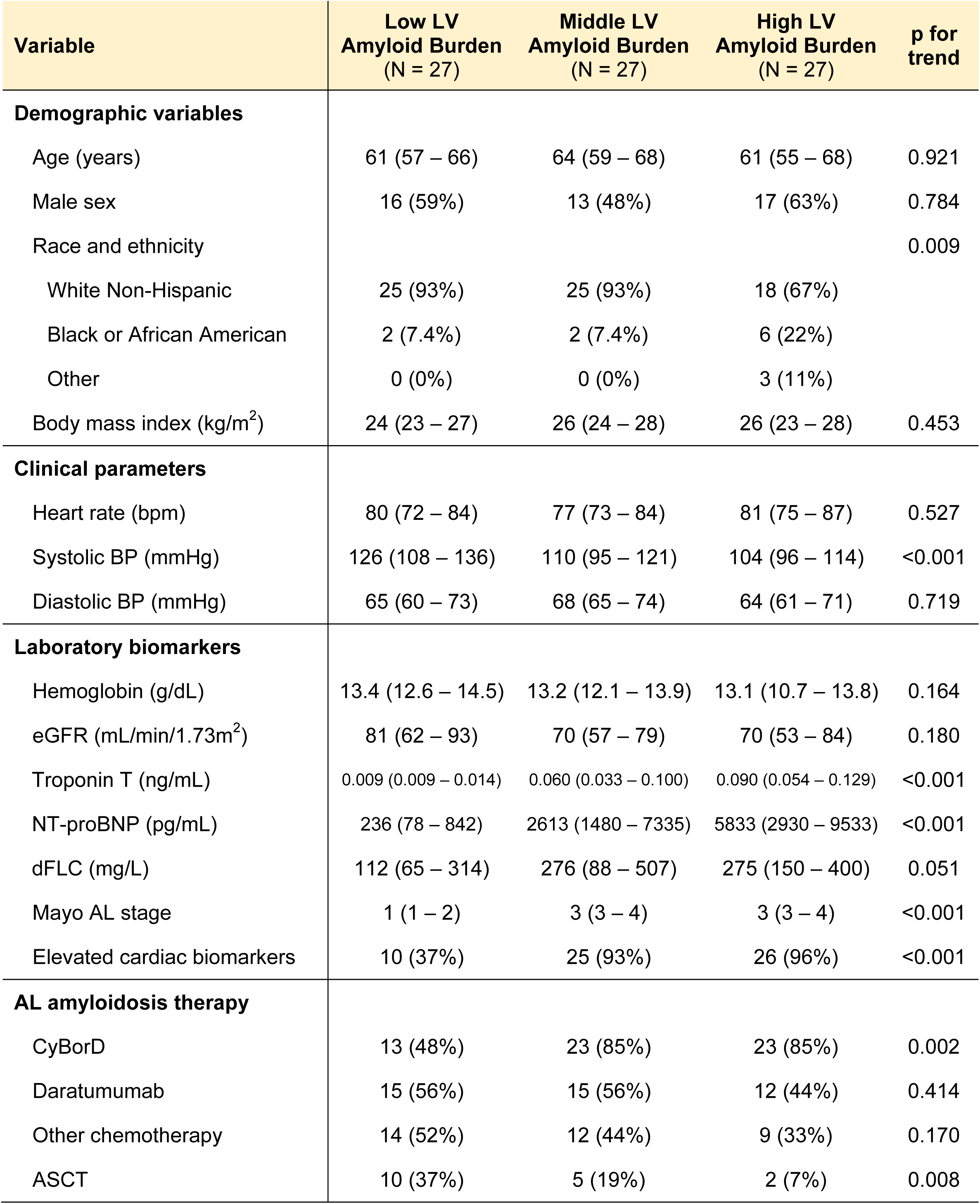
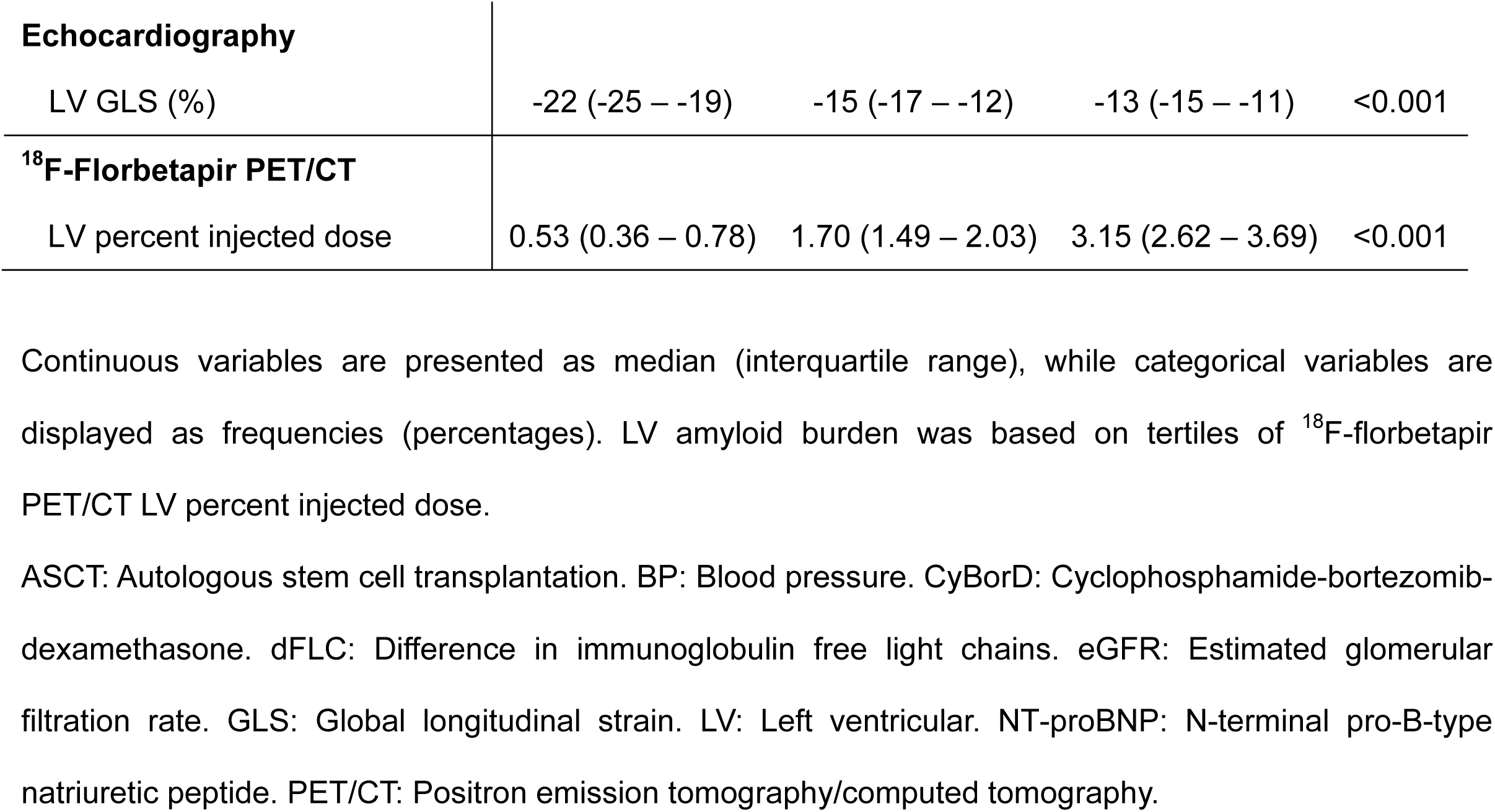
Participant Characteristics at Baseline.

| Variable | Low LV<br>Amyloid Burden<br>(N = 27) | Middle LV<br>Amyloid Burden<br>(N = 27) | High LV<br>Amyloid Burden<br>(N = 27) | p for<br>trend |
| --- | --- | --- | --- | --- |
| **Demographic variables** |  |  |  |  |
| Age (years) | 61 (57 – 66) | 64 (59 – 68) | 61 (55 – 68) | 0.921 |
| Male sex | 16 (59%) | 13 (48%) | 17 (63%) | 0.784 |
| Race and ethnicity |  |  |  | 0.009 |
| White Non-Hispanic | 25 (93%) | 25 (93%) | 18 (67%) |  |
| Black or African American | 2 (7.4%) | 2 (7.4%) | 6 (22%) |  |
| Other | 0 (0%) | 0 (0%) | 3 (11%) |  |
| Body mass index (kg/m<sup>2</sup>) | 24 (23 – 27) | 26 (24 – 28) | 26 (23 – 28) | 0.453 |
| **Clinical parameters** |  |  |  |  |
| Heart rate (bpm) | 80 (72 – 84) | 77 (73 – 84) | 81 (75 – 87) | 0.527 |
| Systolic BP (mmHg) | 126 (108 – 136) | 110 (95 – 121) | 104 (96 – 114) | <0.001 |
| Diastolic BP (mmHg) | 65 (60 – 73) | 68 (65 – 74) | 64 (61 – 71) | 0.719 |
| **Laboratory biomarkers** |  |  |  |  |
| Hemoglobin (g/dL) | 13.4 (12.6 – 14.5) | 13.2 (12.1 – 13.9) | 13.1 (10.7 – 13.8) | 0.164 |
| eGFR (mL/min/1.73m<sup>2</sup>) | 81 (62 – 93) | 70 (57 – 79) | 70 (53 – 84) | 0.180 |
| Troponin T (ng/mL) | 0.009 (0.009 – 0.014) | 0.060 (0.033 – 0.100) | 0.090 (0.054 – 0.129) | <0.001 |
| NT-proBNP (pg/mL) | 236 (78 – 842) | 2613 (1480 – 7335) | 5833 (2930 – 9533) | <0.001 |
| dFLC (mg/L) | 112 (65 – 314) | 276 (88 – 507) | 275 (150 – 400) | 0.051 |
| Mayo AL stage | 1 (1 – 2) | 3 (3 – 4) | 3 (3 – 4) | <0.001 |
| Elevated cardiac biomarkers | 10 (37%) | 25 (93%) | 26 (96%) | <0.001 |
| **AL amyloidosis therapy** |  |  |  |  |
| CyBorD | 13 (48%) | 23 (85%) | 23 (85%) | 0.002 |
| Daratumumab | 15 (56%) | 15 (56%) | 12 (44%) | 0.414 |
| Other chemotherapy | 14 (52%) | 12 (44%) | 9 (33%) | 0.170 |
| ASCT | 10 (37%) | 5 (19%) | 2 (7%) | 0.008 |
| **Echocardiography** |  |  |  |  |
| LV GLS (%) | -22 (-25 – -19) | -15 (-17 – -12) | -13 (-15 – -11) | <0.001 |
| **<sup>18</sup>F-Florbetapir PET/CT** |  |  |  |  |
| LV percent injected dose | 0.53 (0.36 – 0.78) | 1.70 (1.49 – 2.03) | 3.15 (2.62 – 3.69) | <0.001 |

Continuous variables are presented as median (interquartile range), while categorical variables are displayed as frequencies (percentages). LV amyloid burden was based on tertiles of <sup>18</sup>F-florbetapir PET/CT LV percent injected dose.
ASCT: Autologous stem cell transplantation. BP: Blood pressure. CyBorD: Cyclophosphamide-bortezomib-dexamethasone. dFLC: Difference in immunoglobulin free light chains. eGFR: Estimated glomerular filtration rate. GLS: Global longitudinal strain. LV: Left ventricular. NT-proBNP: N-terminal pro-B-type natriuretic peptide. PET/CT: Positron emission tomography/computed tomography.

### Adverse Outcomes

Within 12 months, 29 participants experienced MACE (36%), among whom 13 died (16%), 20 were hospitalized for heart failure (25%), and 3 underwent cardiac transplantation (4%). MACE, all-cause death and heart failure hospitalization were more frequent with higher LV amyloid burden (all p<0.001; Table 2). During the complete available follow-up (median 26 months, IQR 3–45), 40 participants experienced MACE (49%), and 22 died (27%; Supplemental Table 2).

**Table 2.** Adverse Outcomes Within 12 Months.

| **Event** | **Low LV**<br>**Amyloid Burden**<br>**(N = 27)** | **Middle LV**<br>**Amyloid Burden**<br>**(N = 27)** | **High LV**<br>**Amyloid Burden**<br>**(N = 27)** | **p for**<br>**trend** |
| --- | --- | --- | --- | --- |
| All-cause death | 1 (4%) | 5 (19%) | 7 (26%) | 0.026 |
| Heart failure hospitalization | 1 (4%) | 8 (30%) | 11 (41%) | 0.002 |
| Cardiac transplantation | 0 (0%) | 1 (3.7%) | 2 (7%) | 0.150 |
| MACE | 2 (7%) | 10 (37%) | 17 (63%) | <0.001 |
| Time to first MACE (months) | 12.0 (12.0 – 12.0) | 12.0 (2.1 – 12.0) | 3.9 (0.8 – 12.0) | <0.001 |

Categorical variables are presented as frequencies (percentages), while time to first MACE (or to censoring) is presented as median (interquartile range).

### Kaplan-Meier Analysis of LV Amyloid Burden with MACE and All-Cause Death

Increasing tertiles of LV amyloid burden by ^18^F-florbetapir %ID were significantly associated with both MACE and all-cause death within 12 months (Figure 2). For MACE, we found p<0.001 for trend across tertiles, and pairwise testing showed: low vs. middle tertile p=0.014, middle vs. high tertile p=0.056 and low vs. high tertile p<0.001. For all-cause death, we found p<0.027 for trend across tertiles, and pairwise testing showed: low vs. middle tertile p=0.141, middle vs. high tertile p=0.462 and low vs. high tertile p=0.074. Using log-rank statistic maximization, the optimal LV %ID cut-off was >2.0 to predict both MACE and all-cause death.

**Figure 2.**
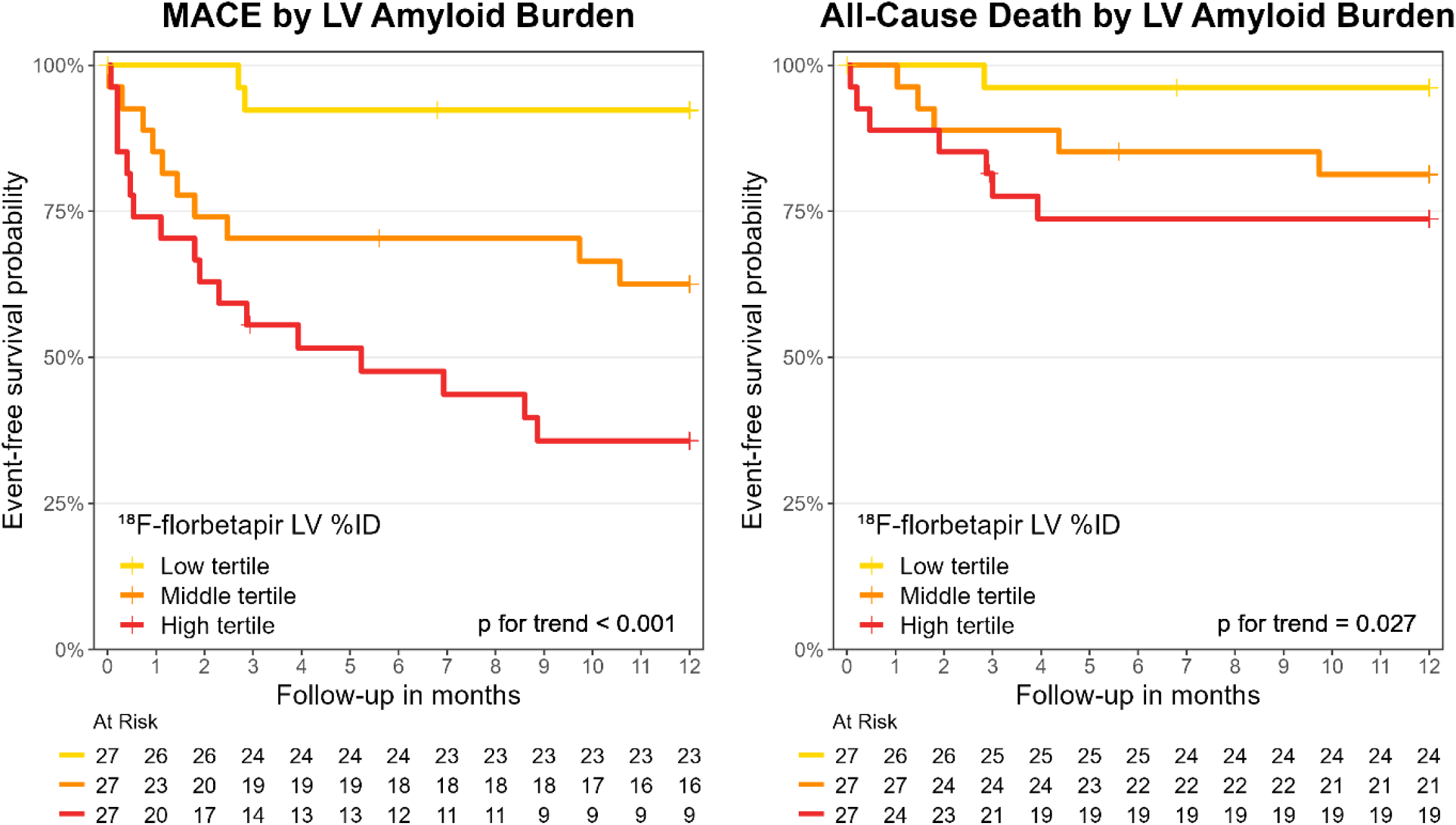
Outcome Analysis Within 12 Months Based on LV Amyloid Burden. Kaplan-Meier analysis and the log-rank test were performed by levels of LV amyloid burden based on tertiles of ^18^F-florbetapir LV %ID. %ID: Percent injected dose. LV: Left ventricular. MACE: Major adverse cardiac events (all-cause death, heart failure hospitalization or cardiac transplantation).

### Univariable Predictors of MACE and All-Cause Death

In Cox regression for outcomes within 12 months, troponin T and NT-proBNP were associated with MACE and all-cause death, but dFLC was only associated with all-cause death, while age and sex were not associated with adverse outcomes (Table 3). In particular, LV amyloid burden by ^18^F-florbetapir %ID was associated with MACE and all-cause death (p=0.002 and p=0.028). ^18^F-florbetapir LV %ID was higher in participants who experienced MACE compared to those who did not: median 2.30 (IQR 1.96–3.08) vs. 1.33 (0.52–2.10, p<0.001). ASCT was associated with lower MACE and chemotherapies other than CyBorD and daratumumab with lower death. These associations may be explained by the initial treatment choice based on predicted risk.

**Table 3.**
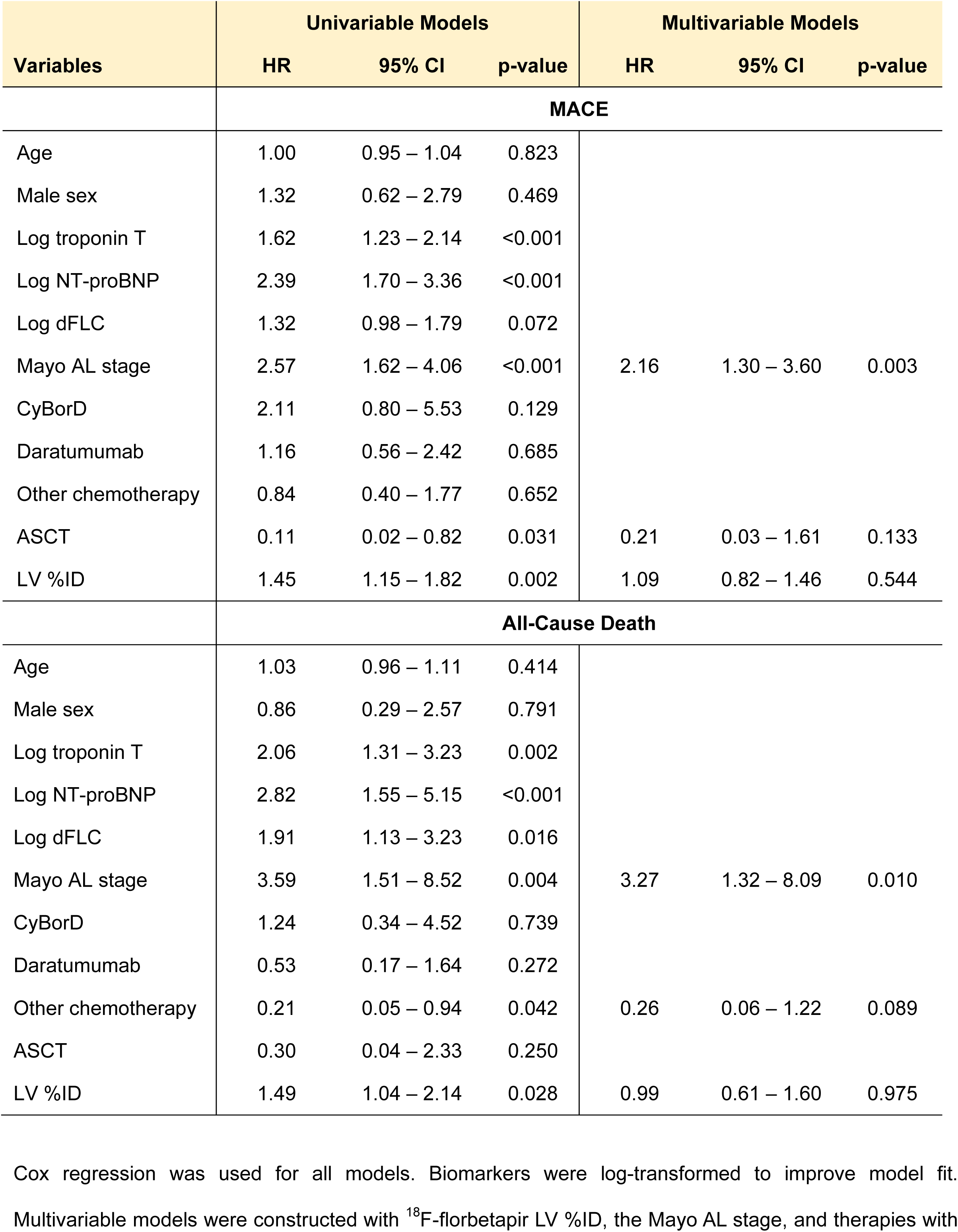
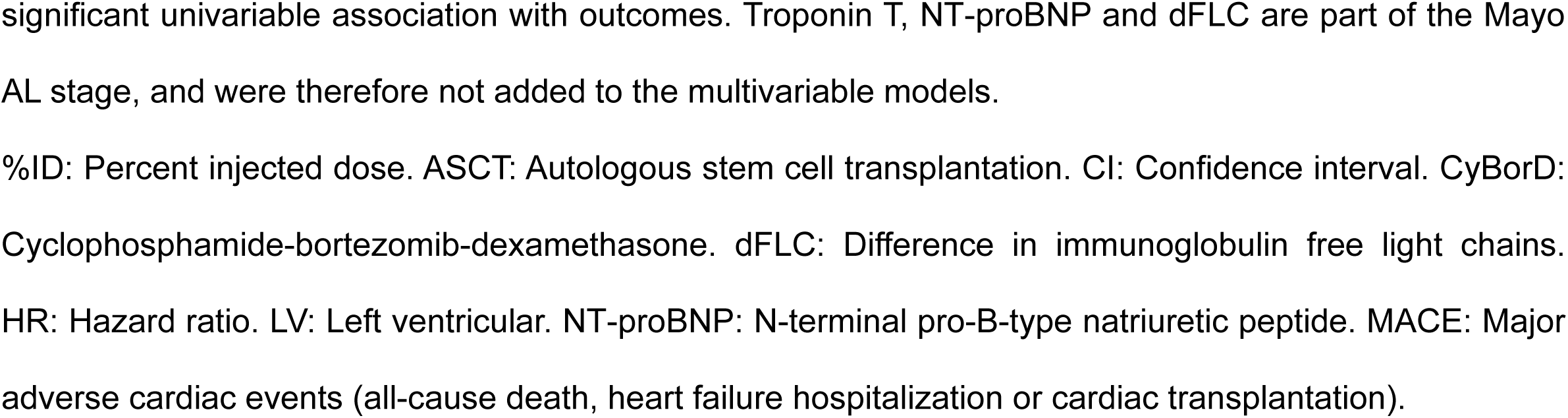
Predictors of Adverse Outcomes Within 12 Months.

### Independent Association of LV Amyloid Burden with MACE or All-Cause Death

In multivariable Cox regression including the Mayo stage, LV amyloid burden by ^18^F-florbetapir %ID did not remain independently associated with MACE or all-cause death (Table 3). A potential explanation is a mediation by the components of the Mayo stage, which was investigated using mediation analysis. Moreover, therapies were no longer associated with outcomes after adjustment for the Mayo stage, which supports the interpretation that the univariable associations were reflecting treatment choice based on the Mayo stage.

### Mediation Analysis of the Relationship Between LV Amyloid Burden and MACE

Our causal mediation model evaluated the magnitude of the relationship between LV amyloid burden (by ^18^F-florbetapir %ID) and MACE, directly or indirectly through cardiomyocyte stretch (by NT-proBNP) or myocardial injury (by troponin T), accounting for confounding by circulating light chains (by dFLC). We found that LV amyloid burden was associated with MACE primarily through an indirect pathway over cardiomyocyte stretch (p<0.001), while the indirect pathway over myocardial injury, and the direct pathway to MACE were statistically non-significant (Figure 3). Circulating light chains showed no significant confounding of the association between LV amyloid burden and MACE. Performing the mediation analysis on all-cause death instead of MACE led to similar results.

**Figure 3.**
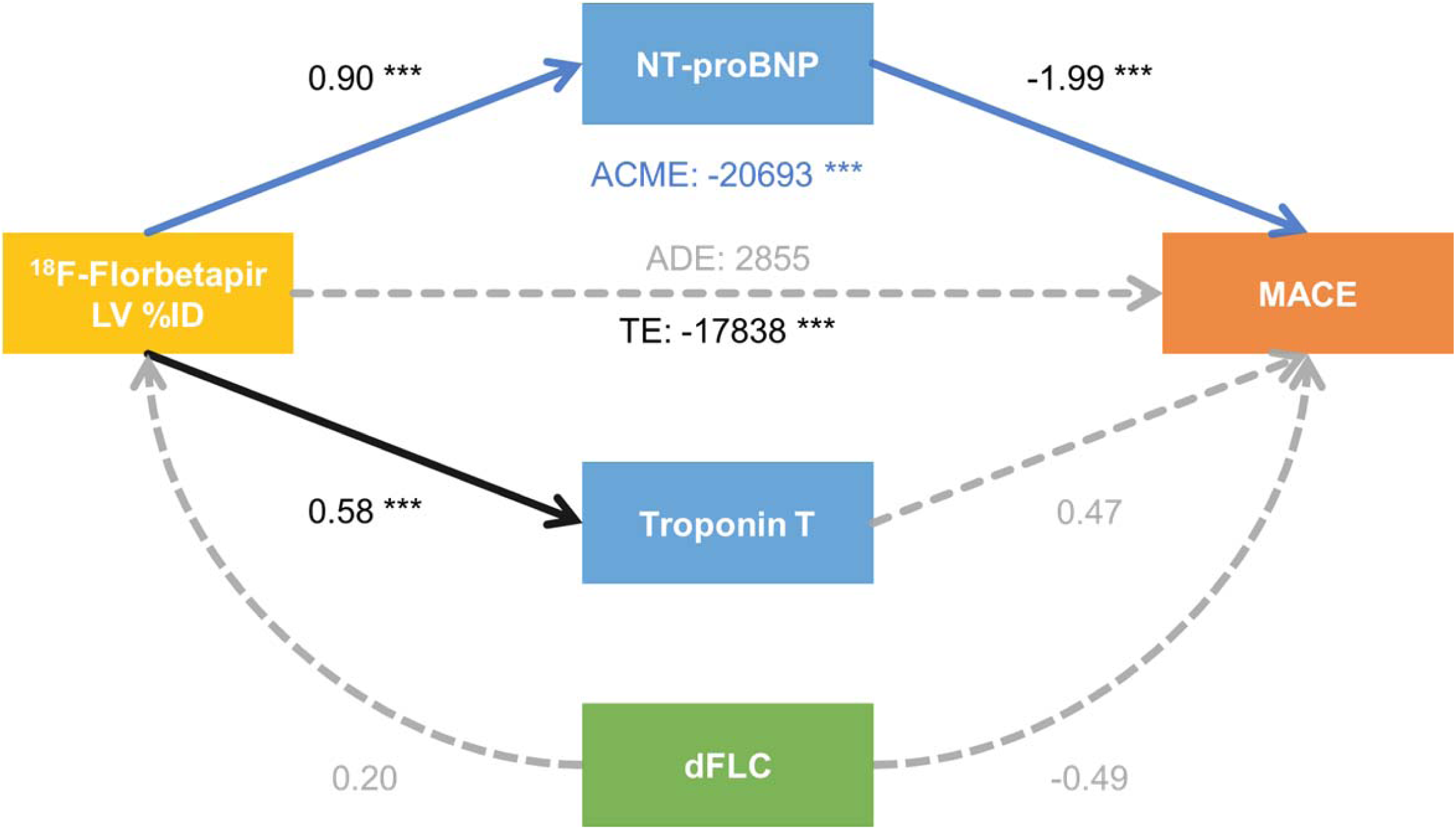
Mediation Analysis Between LV Amyloid Burden and MACE Within 12 Months. This causal mediation model quantifies how LV amyloid burden (by ^18^F-florbetapir LV %ID, yellow) predicts 12-month MACE (orange) directly and indirectly through cardiomyocyte stretch and light chain toxicity (by NT-proBNP as a mediator, blue) and myocardial injury (by troponin T as a mediator, blue), adjusted for circulating light chains (by dFLC as a confounder, green). Arrows represent the direction of assumed causality, with grey dashed arrows for statistically non-significant associations. Results are presented as regression coefficients with * p<0.05, ** p<0.01 and *** p<0.001. LV amyloid burden was associated with MACE (total effect [TE]: -17838, 95%CI -7998529 – -610, p<0.001) essentially through an indirect pathway over cardiomyocyte stretch (NT-proBNP; average causal mediation effect [ACME]: -20693, 95%CI -12877245 – -667, p<0.001), while the indirect pathway over myocardial injury (troponin T), the direct pathway to MACE (average direct effect [ADE]: 2855, 95%CI -101307 – 2073952, p=0.590), and the confounding by circulating light chains (dFLC) were non-significant. %ID: Percent injected dose. CI: Confidence interval. dFLC: Difference in immunoglobulin free light chains. LV: Left ventricular. MACE: Major adverse cardiac events (all-cause death, heart failure hospitalization or cardiac transplantation). NT-proBNP: N-terminal pro-B-type natriuretic peptide.

### Sensitivity Analyses

Using endpoints during the complete available follow-up, outcome analyses and mediation analyses led to identical conclusions (<u>Supplemental Materials</u>).

## Discussion

This prospective study of 81 participants with newly-diagnosed systemic AL amyloidosis revealed three key novel findings. First, LV amyloid burden quantified by LV ^18^F-florbetapir %ID on PET/CT is a significant predictor of MACE and all-cause death. Second, adjustment for the Mayo AL stage, based on troponin T, NT-proBNP and dFLC, attenuated the association of ^18^F-florbetapir LV %ID with MACE and all-cause death. Third, mediation analysis implied that the association between ^18^F-florbetapir LV %ID and MACE is primarily mediated by NT-proBNP, which reflects cardiomyocyte stretch and light chain toxicity, and quantifies heart failure severity. Thus, amyloid burden contributes to adverse outcomes in AL amyloidosis via cardiomyocyte stretch, light chain toxicity and heart failure. This mediation may explain why the adjustment for the Mayo AL stage made the association between ^18^F-florbetapir LV %ID and outcomes non-significant.

In AL amyloidosis, echocardiographic strain and diastolic metrics, as well as late gadolinium enhancement, T1 and extracellular volume on cardiac MRI, are known to predict adverse outcomes.^8–11^ In PET, previous studies have only reported that adverse outcomes were associated with ^11^C-Pittsburgh compound B cardiac uptake, measured either as visually positive, or as ratio of maximal LV uptake / mean aortic uptake.^19,20^ The present study demonstrates for the first time that amyloid burden estimated by ^18^F-florbetapir LV %ID predicts adverse outcomes. Furthermore, we observed more severe alterations in cardiac biomarkers, as well as structural and functional metrics from echocardiography, in participants with higher LV amyloid burden. Together, these findings support the ability of ^18^F-florbetapir PET/CT with LV %ID to quantify LV amyloid burden and the severity of cardiac amyloidosis, which predicts adverse outcomes. This capability is essential to the development of ^18^F-florbetapir PET/CT as a potential surrogate biomarker that could accelerate the development of novel fibril-depleting therapies to reduce amyloid burden.^31,32^

Most prior studies evaluating the prognostic value of cardiac imaging measures did not include adjustment for Mayo AL stage. We have shown that the association between amyloid burden and MACE became non-significant when adjusted for the Mayo AL stage. We hypothesized that this may be due to mediation by one or more of the components of the Mayo AL stage: troponin T, NT-proBNP, and dFLC. Therefore, we performed a mediation analysis based on these biomarkers, which generated new mechanistic insights into the association between LV amyloid burden and adverse outcomes. Our findings suggest that this association is primarily mediated by NT-proBNP, a marker of cardiomyocyte stretch and light chain toxicity. Therefore, it appears that the mere presence of amyloid in the heart does not lead to adverse outcomes. Rather, amyloid burden substantial enough to cause mechanical stress and ultimately heart failure is necessary to lead to adverse outcomes in AL amyloidosis. This finding may be intuitive, but it has not previously been demonstrated. Indeed, until now, there was no specific and quantitative tool to assess amyloid burden. This is now available with amyloid-binding PET radiotracers. In our study, troponin T was not an independent predictor of outcomes, unlike in previous works, which found a significant prognostic value.^5^ Thus, we did not find a significant mediation pathway through troponin T as a marker of myocardial injury. Based on our data, we conclude that the cardiomyocyte stretch pathway predominates over the myocardial injury pathway and over a direct pathway. These pathophysiologic insights on the consequences of amyloid deposition in the myocardium complement previous research on the toxic effects of circulating light chains on cardiomyocyte function.^33,34^

This study had limitations. First, the small sample size limited our statistical power to analyze associations with outcomes. However, our statistically significant results support important effect sizes, thus clinically relevant findings. Moreover, our cohort is among the largest for systemic AL amyloidosis. Second, hematological response was unknown in participants experiencing early death, and almost every participant surviving up to 12 months had a favorable hematological response. Therefore, hematological response could not be meaningfully included in survival models. However, our study also had strengths, such as the prospective, structured cohort design, the inclusion of AL participants with and without cardiomyopathy, all of them with new diagnosis, the quantitative evaluation of LV amyloid burden by ^18^F-florbetapir PET/CT, the comparison with the Mayo stage, and the mediation analysis.

In conclusion, LV amyloid burden quantified by ^18^F-florbetapir LV %ID in PET/CT is associated with worse cardiac function and worse cardiac biomarkers, and predicts adverse outcomes in participants with systemic AL amyloidosis, albeit not independent of the Mayo AL stage. Through mediation analysis, we gained novel pathophysiological insights suggesting that the impact of LV amyloid burden on adverse outcome is mediated by NT-pro-BNP, a marker of cardiomyocyte stretch, light chain toxicity, and heart failure. Together, these results establish quantitative ^18^F-florbetapir PET/CT as a valid method to predict adverse outcomes in AL amyloidosis. This method could become pivotal as an amyloid-specific surrogate outcome in future clinical trials on novel therapies for systemic AL amyloidosis.

## Data Availability

The data underlying this article cannot be shared publicly due to data privacy as defined in the informed consent document.

## Acknowledgments

We are extremely grateful to each of the study participants and their families for their participation and to our funding partners for making this study possible.

## Sources of funding

This work was supported by the National Institutes of Health.

Dorbala: R01 HL 130563; K24 HL 157648; AHA16 CSA 2888 0004; AHA19SRG34950011

Falk: R01 HL 130563

Liao: AHA16 CSA 2888 0004; AHA19SRG34950011

Ruberg: R01 HL 130563; R01 HL 093148

https://clinicaltrials.gov/ct2/show/NCT02641145

## Abbreviations

%ID: Percent injected dose
AL: Light-chain amyloidosis
CT: Computed tomography
dFLC: Difference between involved and uninvolved immunoglobulin free light chains
LV: Left ventricular
MACE: Major adverse cardiac events
MICA: Molecular Imaging of Primary Amyloid Cardiomyopathy Study
NT-proBNP: N-terminal pro-B-type natriuretic peptide
PET: Positron emission tomography
VOI: Volume of interest

## Graphical Abstract

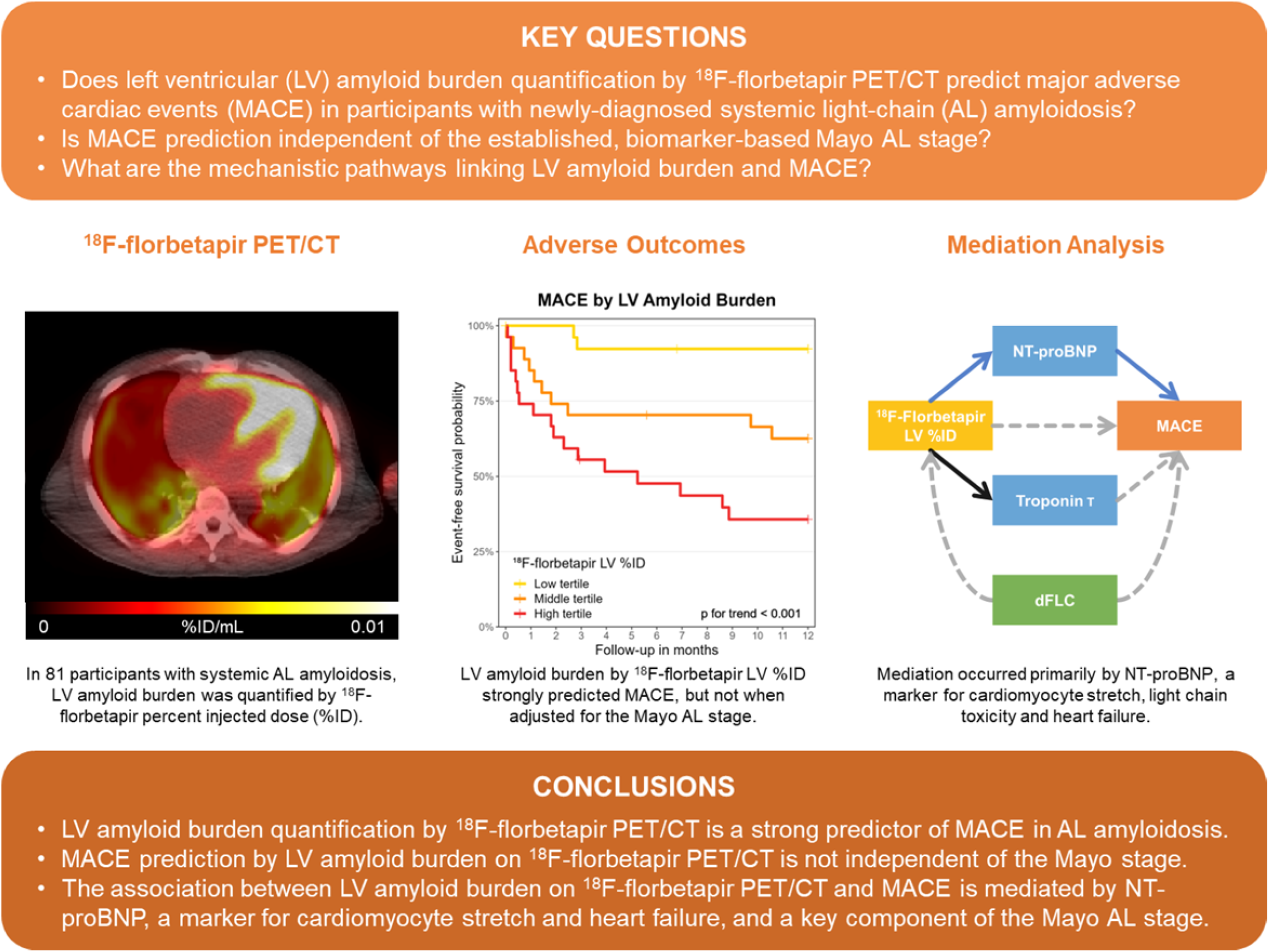

